# Genetic insights into smoking behaviours in 10,558 men of African ancestry from continental Africa and the UK

**DOI:** 10.1101/2021.12.09.21267119

**Authors:** Noemi-Nicole Piga, Palwende Romuald Boua, Chisom Soremekun, Nick Shrine, Kayesha Coley, Jean-Tristan Brandenburg, Martin D. Tobin, Michèle Ramsay, Segun Fatumo, Ananyo Choudhury, Chiara Batini

## Abstract

Smoking is a leading risk factor for many of the top ten causes of death worldwide. Of the 1 billion smokers globally, 80% live in low- and middle-income countries, where the number of deaths due to tobacco use is expected to double in the next decade according to the World Health Organization. Genetic studies have helped to identify biological pathways for smoking behaviours, but have mostly focussed on individuals of European ancestry or living in either North America or Europe.

Here we present a genome-wide association study of two smoking behaviour traits in 10,558 men of African ancestry living in five African countries and the UK. Eight independent variants were associated with either smoking initiation or cessation at p-value < 5 × 10^−6^. Of these, four were monomorphic or rare in European populations. Gene prioritization strategy highlighted five genes, including *SEMA6D*, previously described as associated with several smoking behaviour traits. These results confirm the importance of genetic epidemiological studies in underrepresented populations.

## Introduction

Smoking is a leading risk factor for many of the top ten causes of death worldwide, including heart and lung diseases^1^. Each year, tobacco use is directly responsible for 7 million deaths and 25% of all cancer fatalities globally^2^. However, smoking prevalence varies among world regions, and of the 1.3 billion tobacco users worldwide, 80% live in low- and middle-income countries (LMICs)^3^.

Reassuringly, prediction models by the World Health Organization (WHO) show a reduction in smoking prevalence in most areas from 2010 to 2025 thanks to tobacco control strategies^4,5^. However, in parallel to the American and European decline, the tobacco industry and market has seen a recent expansion in the African continent, due to the fast population growth and the improvement in buying power^6^. In line with this, sub-Saharan Africa has experienced a 52% increase in tobacco use from 1980 to 2016^7^ and a further 9 million people are expected to take up smoking in the African region by 2025^8^. In addition, the support for smokers wishing to quit and for healthcare professionals training in smoking cessation is still very limited across the continent^9^. To help counteract these changes, as of November 2018 more than forty African countries supported the WHO Framework Convention on Tobacco Control and twenty countries were involved in the Protocol to Eliminate Illicit Trade in Tobacco Products^6^.

Currently, there are 94 million male and 13 million female tobacco users in Africa, with one in five adolescents using tobacco, and an increasing smoking prevalence among young women^6^. The type of tobacco used varies among countries and sexes, and its consumption is overall associated with alcohol drinking, lower income status, education levels, and professional activity^10,11^. Using Demographic Health Surveys data for 30 sub-Saharan African countries, Sreeramareddy et al. compared smoking with the use of smokeless tobacco and showed that while the first is more prevalent among men, the majority of women preferentially uses the second, and specifically chewing tobacco^10^. Similar patterns were confirmed by a recent study focusing on tobacco and alcohol use in rural and urban settings in four African countries^11^.

The patterns described above change when we focus on communities of African descent in the UK. In 2019, 14% of the adult UK general population smoked regularly, with slightly different figures between sexes: 15.9% of men and 12.5% of women smoked, resulting in a ratio between male and female smoking proportions of 1.27. This ratio was 1.87 among Black ethnic minorities in the same year^12^; however, contrary to what is observed in continental Africa, between 2014 and 2019 the smoking prevalence among Black adults decreased from 13.3% to 9.6%^12^.

In the US, African-American individuals have been shown, on average, to initiate smoking later and smoke fewer cigarettes per day than European-American individuals^13^. However, they show comparable levels of nicotine equivalents, are less likely to successfully quit smoking, and have a higher risk of smoking-related lung cancer^13,14^.

Genetics has been shown to play a role in smoking behaviour traits. Genome-wide association studies (GWASs) identify genetic variants associated with the trait of interest, which inform biological understanding and highlight functional pathways and potential drug targets for precision medicine approaches to treatment^15^. The strongest associations with smoking behaviours have been consistently shown at locus 15q25.1 for amount smoked and nicotine dependence, containing the cluster of *CHRNA5-A3-B4* genes which encode subunits of the nicotinic acetylcholine receptors (nAChRs)^16^. In the brain, nicotine binds to nAChRs stimulating the release of several neurotransmitters, impacting the reward pathway, learning and memory^16^. To date, the largest GWAS of smoking behaviour traits includes 1.2 million individuals and highlights genes coding for proteins involved in neurotransmission of nicotine, dopamine and glutamate^17^. However, as in most GWASs, it only includes individuals of European origin failing on the representation of global human diversity^18,19^.

Previous genetic epidemiology studies of smoking behaviour in individuals of African descent have only included African-American (AA) participants and have identified four genetic loci associated with different traits. Located ∼6kb from the 5’ of *CHRNA5*, rs2036527 on chromosome (chr) 15, was the only variant associated with cigarettes per day in 32,389 AA individuals in the Study of Tobacco in Minority Populations^13^. In a study including ∼1000 AA participants, Chenoweth et al. found three independent variants on chr 19 (rs12459249, rs111645190, rs185430475) associated with nicotine metabolite ratio (NMR)^14^. These variants showed low linkage disequilibrium with four signals previously identified in a Finnish cohort and located in the genomic region of the *CYP2A6* gene which encodes for the key enzyme of nicotine metabolism^14,16^. Hancock et al. performed a trans-ethnic analysis including 28,677 European and 9,925 AA smokers, and identified variant rs910083 on chr 20 as associated with nicotine dependence^20^. Finally, Xu et al. studied smoking trajectories in almost 300,000 individuals from the Million Veteran Program (MVP), including >54,000 AAs, which aimed to capture the variation of smoking status over time^21^. They found an association on chr 1 with variant rs4478781 in AAs only, and 14 associated loci in a trans-ethnic meta-analysis including European-Americans and Hispanic-Americans, mainly driven by the results from the European ancestry group^21^.

Despite these interesting findings, individuals of African descent, and especially those living in continental Africa, remain heavily unrepresented in large-scale studies^18,19^. Here we present the first GWAS of smoking behaviour traits in 10,558 men of African ancestry living in five African countries and the UK. In agreement with the sex biases previously described^10,11^, for the African datasets included here, smoking prevalence in females ranged between 0.5% and 7%, which prompted their exclusion from all discovery analyses.

## Materials and methods

### Participants

This study used genetic data and smoking behaviour information from three different cohorts: the Africa Wits-INDEPTH Partnership for Genomic Studies (AWI-Gen)^22^, the Uganda Genome Resource^23^(UGR) and the individuals of African ancestry in UK Biobank (UKB-AFR)^24^. AWI-Gen is a cross-sectional population study including ∼12,000 individuals from Ghana, Burkina Faso, Kenya and South Africa, aged 40-60+ years, aimed at understanding the genomic and environmental factors that contribute to body composition and cardio-metabolic diseases. For the purpose of this study, with the aim of accounting for the local population structure, the AWI-Gen cohort was divided into three datasets representing the main geographical areas: AWI-East (Kenya), AWI-South (South Africa) and AWI-West (Ghana and Burkina Faso).

The UGR includes ∼6,400 individuals, and it is a subset of the General Population Cohort (GPC)^25^, a population-based open cohort aimed at understanding HIV infections in Uganda. The UGR was built with the aim of improving the local resources for public health and to allow large genetic epidemiology studies^23^.

UKB is a large longitudinal study in the UK which includes samples from over 500,000 volunteers aged 40-69 years at baseline^24^. Since 2006, it has collected extensive phenotypic and biological data in order to allow approved researchers to investigate the genetic and/or environmental determinants of a wide range of diseases and health-related phenotypes. Using genetic data for population stratification, we identified ∼7,800 individuals of African ancestry in UKB^26^.

### Description of phenotypes

We have analysed two binary smoking behaviour traits: Smoking Initiation (SI) and Smoking Cessation (SC), defined using relevant answers in questionnaire data available in each cohort. Overall, SI compares ‘never’ smokers (controls) versus ‘ever’ smokers (cases), in which the former represents individuals who have never, or only very rarely, smoked in their life and the latter are participants who smoked or currently smoke every day or occasionally. SC only includes ‘ever’ smokers and it compares ‘current’ smokers (cases), who were smoking every day or occasionally at the time of answering the questionnaire, to ‘previous’ smokers (controls), who were not. The detailed description of the phenotype definitions and the specific questions used for each cohort are available in Supplementary Note 1.

The number of cases and controls for each phenotype in each cohort is presented in Table 1.

**Table 1.** Sample size (and mean age) for each phenotype in each dataset for cases (1) and controls (0).

| Cohort | SI - Smoking initiation<br>ever (1) vs never (0) |  | SC - Smoking cessation<br>current (1) vs previous (0) |  |
| --- | --- | --- | --- | --- |
|  | ever | never | current | previous |
| AWI-east | 425<br>(50yrs) | 374<br>(49yrs) | 185<br>(49yrs) | 239<br>(50yrs) |
| AWI-south | 1343<br>(53yrs) | 811<br>(56yrs) | 876<br>(51yrs) | 466<br>(56yrs) |
| AWI-west | 782<br>(50yrs) | 1045<br>(50yrs) | 481<br>(50yrs) | 301<br>(51yrs) |
| UGR | 544<br>(50yrs) | 2100<br>(27yrs) | 420<br>(49yrs) | 124<br>(57yrs) |
| UKB-AFR | 1167<br>(52yrs) | 1967<br>(51yrs) | 509<br>(50yrs) | 656<br>(54yrs) |

### Genotyping and Imputation Quality Control

The AWI-Gen individuals were genotyped at ∼2.4M SNPs using the Illumina Infinium H3Africa SNP array, which is designed to be specific and sensitive to the genomic diversity of African populations^22^. Imputation at ∼39M autosomal variants was performed using the Haplotype Reference Consortium (HRC) panel^27^ on the Michigan Imputation Server. Details of quality control (QC) and imputation settings are presented in Choudhury et al al. 2020^28^. The UGR individuals were genotyped at ∼2.2M autosomal markers using the HumanOmni2.5-8 chip array. Imputation at ∼98M variants was performed using a combined reference panel with sequence data from three different studies (African Genome Variation Project^29^, Uganda 2000 Genomes and 1000 Genomes Project Phase 3 [1000GP]^30^). Details of QC and imputation settings are presented in Gurdasani et al.^23^.

UK Biobank individuals were genotyped at ∼800,000 variants using the UK Biobank Axiom Array. Imputation at ∼93M autosomal variants was performed using the HRC^27^, the UK10K^31^ and the 1000GP^30^ reference panels combined. Details of QC and imputation settings are presented in Bycroft et al.^24^.

Details of genotyping QC are reported in Supplementary Table 1. Additional QC was performed across all datasets to ensure that effect alleles were consistently aligned between cohorts. Imputed autosomal variants with a minor allele count (MAC) >= 20 or minor allele frequency (MAF) > 0.01, and an imputation info score >= 0.3 were included in all further analyses (Supplementary Figure 1).

### Study level genome-wide association analyses

Genome-wide association analyses were performed using a univariate linear mixed model (LMM) and significance was evaluated using a likelihood ratio test as implemented in GEMMA v0.98.1^32^. Covariates included age, age^2^ and as many principal components (PCs) as required in each dataset (east 5PCs; south 14PCs;west 11PCs; UGR 10PCs; UKB 9PCs). PCs were calculated for each dataset from a PC analysis using independent genotyped SNPs in PLINK v1.90^33^. For AWI-Gen and UKB-AFR datasets, we determined the number of PCs to include by using the eigenvalues to assess when adding further components would not contribute additional information. In an iterative process, we stopped at the first PC for which the contribution of the three previous PCs was not greater than the contribution of the following three. For UGR, 10 PCs were used as in Gurdasani et al.^23^. The genetic relatedness matrix (GRM) included in the LMM was calculated for each full cohort with GEMMA v0.98.1^32^ using independent SNPs. Details of the QC used for genotyped variants and the number of variants used to perform the PCA and to calculate the GRM for each cohort are reported in Supplementary Table 1. Manhattan and quantile-quantile (qq-) plots were visualized using the qqman package in R^34^. The LD score regression intercept was calculated to assess the presence of genomic inflation using ldsc v1.0.1^35^. LD scores were calculated including the African superpopulation from 1000GP^30^. When the LD score intercept was above 1.05, the association p-value was recalculated as follows:

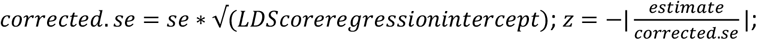

*corrected*.*pvalue* = *P*(*Z* < −*z*) + *P*(*Z* > *z*). Where *corrected. se*and *se* are the corrected and original standard error of the estimate effect size respectively; *z* is the z-score used to calculate the corrected p-value using two-sided test statistics.

### Meta-analysis

We performed a two-step meta-analysis. In step1, we included the three AWI-Gen datasets (AWI-East, AWI-South, AWI-West) to obtain AWI-Gen cohort-level summary statistics. In step2, we included AWI-Gen, UGR and UKB-AFR. In each step, we used the modified random effect model (RE2) as implemented in METASOFT^36^. Variants were included in the analysis if they were present in at least two out of three studies. Supplementary Figure 1 shows the number of variants for each meta-analysis and phenotype. Results were visualized using Manhattan and qq-plots, and LD score regression intercept was calculated to evaluate genomic inflation, as done for the study-level association analyses.

### Definition of associated and sentinel variants

For each trait, variants were divided into three tiers defined using different significance thresholds which take into account the results of the meta-analysis and the cohort-level summary statistics (AWI-Gen, UGR and UKB-AFR). Tier1 included variants with meta-analysis p-value < 5 × 10^−8^ and p-value < 0.05 in each cohort; tier2, variants with a meta-analysis p-value < 5 × 10^−6^ and p-value < 0.05 in each cohort; tier3, variants with a p-value < 5 × 10^−8^ in at least one of the cohort genome-wide association analyses.

For each tier, sentinel variants were defined in an iterative process as the variants with the lowest p-value in a region of 200kb centered on the variant.

### Conditional analysis

To assess the presence of any additional independent signals, we utilized the GCTA v1.93.2^37^ stepwise model selection for the conditional analysis (option --vcojo-slct) in a region of +/- 100kb from each sentinel variant and using the populations of African ancestry in 1000GP^30^ as a reference for LD patterns^30^.

### Fine-mapping analysis

Regions of +/- 100kb from each sentinel variant were analysed to retrieve the 99% credible set variants using FINEMAP v1.4 software^38^ with a shotgun stochastic algorithm assuming one causal variant.

We then created the refined credible sets by filtering for a Posterior Inclusion Probability (PIP) greater than or equal to 1%. LD proxy variants of these SNPs were identified in the African populations in 1000GP^30^ within the regions defined by the 99% credible sets extended by +/-100kb using PLINK v1.90^33^. Variants with both D’ >= 0.9 and r^2^ >= 0.6 were retained. Supplementary Figure 1 and Supplementary Table 2 show the 99% credible sets, the refined credible sets and the proxy variants for each locus. The refined credible set variants and their proxies were used for all follow up analyses.

### Replication and lookup analyses

The replication of our meta-analysis step2 results was performed using two publicly available datasets: (a) the genome-wide summary statistics for smoking trajectories in individuals of African ancestry included in the MVP^21^; (b) and the genome-wide summary statistics from the SI and SC meta-analyses in individuals of European ancestry released by the GWAS & Sequencing Consortium of Alcohol and Nicotine use (GSCAN)^17^. The number of independent sentinel variants was used to calculate the Bonferroni corrected p-value threshold for the replication analyses (0.007 SI; 0.05 SC).

We performed two lookup analyses aiming to understand if our discovered loci were previously described as associated with any smoking phenotypes or any other trait. For the first analysis, we compiled a list of variants described in 14 studies^13,17,20,21,39,40,41,42,43,44,45,46,47,48^ as associated with 9 smoking behaviour traits (Smoking Initiation; Smoking Cessation; Age of Initiation; Cigarettes per Day; Fagerström Test for Nicotine Dependence; Pack Years; Trajectory Contrast I; Trajectory Contrast II; Time to the First Cigarette). For the second analysis, we queried GWAS Catalog v1.0.2^49^ (see URLs) to identify variants associated with any other phenotype.

### Follow up analyses

#### Gene prioritisation

We combined results from four analyses in order to identify the genes influenced by the SNPs in the refined credible sets and their proxies. First, we assessed the predicted pathogenicity of the variants using the Combined Annotation Dependent Depletion (CADD) score^50^ as implemented in the Ensembl Variant Effect Predictor (VEP)^51^. We defined as pathogenic those variants with a CADD score greater or equal than 15.

We then investigated if the variants influenced the expression of a gene or the protein level using eQTL and pQTL data respectively. We used the eQTL Mapping option in the Functional Mapping and Annotation of Genome-Wide Association Studies (FUMA) v1.3.6^52^ which collects eQTL data from 14 data sources (see URLs). Significant eQTLs are defined on either p-value or FDR thresholds based on the specific data source (see URLs) and we retrieved results from blood, brain or lung tissues. For the pQTL analysis, we used data from 90 cardiovascular genes of the SCALLOP Consortium^53^ and followed their definition of significant pQTLs. They defined signals more than 1Mb away from the protein gene as *trans*-pQTLs, and signals that were closer than 1Mb as *cis*-pQTL^53^.

Finally, we used the Chromatin Interaction Mapping tool as implemented in FUMA v1.3.6^52^ to gain insights into possible epigenetic properties of the interrogated variants using the blood, brain and lung tissues (see URLs).

We selected those variants that were identified by at least one of the analyses described above and retrieved the list of prioritised genes for SI and SC.

#### Pathway analysis

The prioritised genes were queried to investigate any biological function and implication in smoking behaviour using the webtool Metascape^54^ which performs pathway enrichment and protein-protein interaction analyses combining information from different databases and -omics data using hierarchical clustering^54^. Pathway enrichment is based on an over-representation analysis^54^, while the protein-protein interaction makes use of the MCODE algorithm^55^, which captures densely connected regions in a complex network^54,55^. For both analyses, we used the default option of Metascape and the Entrez Gene ID as gene name. We considered a pathway being enriched if represented by prioritised genes linked to distinct meta-analysis step2 associated loci.

#### PheWAS analysis

Variants in the refined credible set for the locus of chr 15 and passing at least one of the criteria for our gene prioritisation strategy were included in a PheWAS using the PheWAS R package^56^ in three datasets available from the Integrative Epidemiology Unit (IEU) OpenGWAS project (see URLs)^57,58^: (i) ‘IEU analysis of UK Biobank phenotypes‘^59^ (ukb-b, see URLs) and (ii) ‘Neale lab analysis of UK Biobank phenotypes, round 2‘ (ukb-d, see URLs) for European individuals, and (iii) ‘Pan-ancestry genetic analysis of the UK Biobank performed at the Broad Institute‘^60^ (ukb-e, see URLs) for African individuals only. PheWAS results from ukb-b and ukb-d were combined together since they include distinct phenotypes and refer to the same ancestry group. For each ancestry group we filtered for significant associations after applying a Bonferroni correction for each variant based on the number of tested phenotypes.

## Results

### Discovery analyses

We performed a genome-wide association analysis for each dataset and phenotype combination in a total of 10,558 men for SI and 4,257 for SC. A modified random effect model was implemented for both steps of the meta-analysis on variants present in at least two of the individual datasets. Step1 included the three AWI-Gen datasets and step2 meta-analysed the results of step1 with UGR and UKB-AFR (Supplementary Figure 1). Results for the individual studies and meta-analysis step1 are presented in Supplementary Note 2.

For SI, step2 analysed 14,459,454 SNPs: no genome-wide significant variant was observed, while 99 variants passed the suggestive significance threshold (Figure 1a). The qq-plot showed no residual population structure (Supplementary Figure 2a) and the LD score regression intercept was 0.94. For SC, step2 analysed a total of 14,057,868 variants: no SNPs passed the genome-wide significant threshold and 45 SNPs were below the suggestive significance threshold (Figure 1b). The qq-plot showed no residual population structure (Supplementary Figure 2b), confirmed by an LD-score regression intercept value of 0.88. Following our tier criteria and our definition of sentinel variants, we identify (i) no variant in tier1 for either trait; (ii) 7 sentinel variants for SI and one for SC in tier2; and (iii) one variant in tier3 for SI. In the meta-analysis, the 8 sentinel variants from tier2 show low heterogeneity (I^2^) of effect sizes, as well as having a consistent direction of effect among studies (Table 2 and Supplementary Figure 3). We focused our follow up analyses on the sentinel variants in tier2.

**Figure 1.**
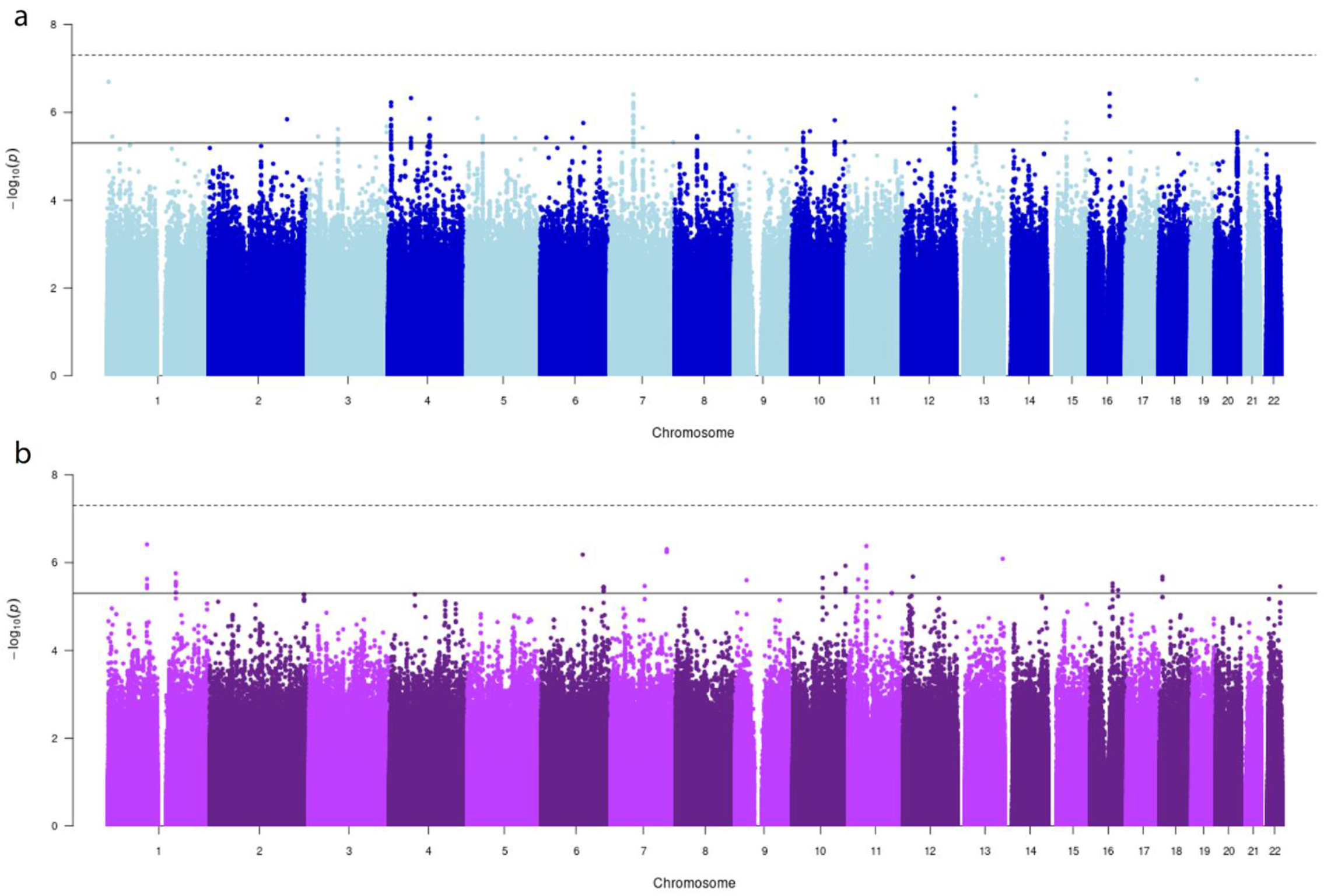
Manhattan plots of step2 GWAS meta-analysis: a) smoking initiation, b) smoking cessation; continuous line, suggestive p-value (p) significance threshold (5 × 10^−6^); dashed line, genome-wide significance threshold (5 × 10^−8^). Number of participants and variants analysed is reported in Supplementary Figure 1.

**Table 2.**
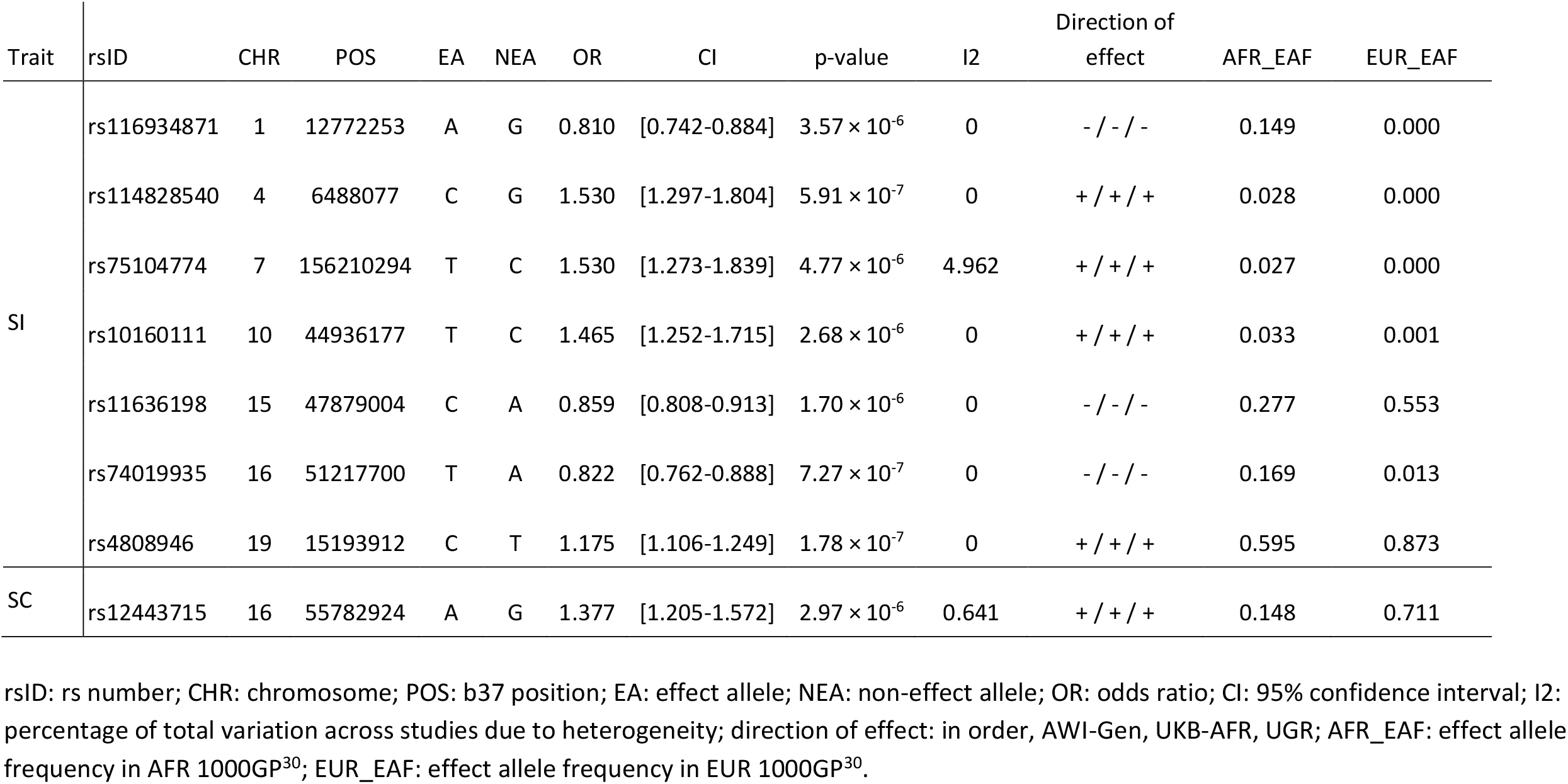
Meta-analysis sentinel variants for smoking initiation (SI) and smoking cessation (SC).

Conditional analyses did not identify any additional independent signal in the 200kb loci we defined around our 8 sentinel variants in tier2. For each locus, we first identified the 99% credible set with a Bayesian approach, which included a total of 2,243 potentially causal variants for SI and 120 for SC (Supplementary Table 2). These 99% credible sets spanned regions of 47.6 to 200kb (see Supplementary Figure 4), with the only exception of the 99% credible set on chr 19 which included only one variant. We then identified the refined credible sets to include only those variants with a PIP > 0.01 and their LD proxies (D’ >= 0.9 and r^2^ >= 0.6) reducing the number of SNPs to 136 for SI and 36 for SC (Supplementary Figure 1 and Supplementary Table 2).

### Replication and literature lookup analyses

We performed replication analyses using the refined credible sets and their proxy variants in two different datasets: (a) an analysis of smoking trajectory contrasts performed in individuals of African ancestry^21^; and (b) two meta-analyses of SI and SC in European individuals^17^. The smoking trajectory contrasts represent a comparison of either (I) current vs never or (II) current vs mixed smokers, defined using electronic health records data, and capturing SI and SC respectively^21^.

When using dataset (a)^21^, 9 out of the 69 variants in the refined credible set on chr 15 passed the Bonferroni corrected threshold for SI (Supplementary Table 3a). Four out of 5 variants on chr 1 passed the nominal significance threshold, and no variants on chr 10 showed evidence of replication. No dataset (a) variants were found for the loci on chr 4, 7, 16 and 19. When using (b)^17^, all the variants in the refined credible set on chr 15 passed the Bonferroni corrected threshold for SI, with four of these being genome-wide significant and having the same direction of effect as in our analysis (Supplementary Table 3b). No variants on chr 7, 16 and 19 showed evidence of replication for SI or SC and no dataset (b) variants were found for the loci on chr 1, 4 and 10.

We investigated if any of the variants included in the credible sets had been previously described as associated with any smoking behaviour trait or any other phenotype. From both analyses, we only obtained results for the SI trait. One variant on chr 15 (rs9646181) was previously described as associated with smoking trajectory contrast I (p-value 4.8 × 10^−10^) (Supplementary Table 3c), in a trans-ethnic meta-analysis including individuals of African, European and Hispanic ancestries and it was mapped as an intronic variant in the gene *SEMA6D*^21^. Looking beyond smoking behaviour traits, we interrogated the GWAS Catalog and found that the variant rs4624724 on chromosome 4 had been previously described as associated with adolescent idiopathic scoliosis (p-value 4 × 10^−8^; Supplementary Table 3d)^61^.

### Functional follow up analyses

#### Gene prioritisation analyses

We combined the results of four different analyses: variant annotation with VEP and its CADD score estimate, eQTL and pQTL mapping, and chromatin interaction mapping. For both traits, all queried variants were annotated as non-coding, or as part of upstream or downstream regulatory regions (VEP^51^) and no variants were identified as a pQTL using the 90 cardiovascular genes of the SCALLOP Consortium^53^.

For the SI trait, we retrieved a total of 95 genes. Specifically, the CADD scores highlighted a total of six possible deleterious variants affecting two genes, *RP11-552E10*.*1* and *SEMA6D* (Supplementary Table 4a). The eQTL mapping showed that 6 variants affect the expression of 4 genes (*AC073133*.*1, FBN1, MAN2B2* and *SLC1A6*) in different brain tissue datasets (BRAINEAC (see URLs), Common Mind Consortium^62^, eQTLGen^63^, PsychENCODE^64^; Supplementary Table 4a). Finally, the chromatin interaction analysis highlighted 42 unique variants, at least one for each associated locus, having an effect on a total of 92 genes in either brain tissues or lung fibroblast cells^64,65,66^ (Supplementary Table 4a). Three of the 95 genes identified were mapped by two of the prioritisation analyses (*AC073133*.*1, MAN2B2*, and *SEMA6D*; Supplementary Table 4a). *AC073133*.*1* showed significant results for eQTLs for brain tissue with one variant (rs6969023) and for chromatin interaction in lung fibroblasts mediated by three variants (rs116530211,rs76374118,rs79338905; Supplementary Table 4a). *MAN2B2* was identified by variant rs73207830 as being an eQTL in blood, and variant rs116755844 indicated chromatin interaction in the Promoter anchored Hi-C loops data from PsychENCODE^64^ (Supplementary Table 4a). *SEMA6D* showed evidence of both pathogenicity based on a high CADD score and chromatin interaction in lung fibroblast cells (Supplementary Table 4a).

For the SC trait, only one variant had a CADD score higher than or equal to 15 but it was annotated as intergenic (data not shown). The combination of the eQTL and the chromatin interaction mapping defined a total of 30 genes associated with 18 distinct variants (Supplementary Table 4b). The eQTL mapping identified two genes, *CES1* and *LPCAT2*. While *LPCAT2* was highlighted by only one dataset in blood (eQTLGen^63^), *CES1* was retrieved by six datasets (BIOSQTL^67^, DICE^68^, eQLTCatalogue^69^, eQTLGen^63^, GTExv8^70^, PsychENCODE^64^) including several blood cell types (B cells, monocytes, and T cells), and lung and brain tissues (Supplementary Table 4b). Both genes showed significant chromatin interaction values discovered in IMR90, a lung fibroblast cell line^65^ (Supplementary Table 4b). The remaining 28 genes showed SNP-mediated chromatin interaction both in the IMR90 cell line^65^ and in the Promoter anchored Hi-C loops data from PsychENCODE^64^ (Supplementary Table 4b).

#### Pathway analysis

We performed a pathway analysis to investigate biological interactions between the prioritised genes using the web-based tool Metascape developed for overrepresentation analysis of genes in biological pathways and protein-protein interaction^54^. We decided to focus only on pathways enriched with genes implicated by different loci, and so we performed this analysis only on the genes prioritised for SI. Fifty-five out of the ninety-five genes had an Entrez Gene ID and were analysed by Metascape resulting in two enriched pathways from Gene Ontology (GO) Resource. ‘Metanephros development’, the process to form the definitive kidney (GO:0001656), was enriched for *FBN1, SHH* and *WFS1* genes (Log(p-value): -3.50; Supplementary Table 5); the ‘developmental growth involved in morphogenesis’ (GO:0060560), a large GeneOntology category including several classes of morphogenesis activities was enriched for *PDPN, SALL1, SEMA6D* and *SHH* genes (Log(p-value): -2.25; Supplementary Table 5). The protein-protein interaction network analysis identified two interactions: *CYP4F3* with *CYP4F8*, and *FBN1* with *WFS1*.

#### PheWAS analysis

We selected variants in the locus on chr 15 for a PheWAS analysis, as this was the only locus to replicate and it harboured *SEMA6D*, which was previously identified by other studies on smoking behaviour traits (see Discussion). We limited the PheWASs to variants supported by at least one of the four criteria of our gene prioritisation analysis obtaining a total of 4 variants: rs11634974, rs11636198, rs12905212, and rs7273389. The number of tested phenotypes for European individuals (ukb-d and ukb-d) varied among variants: from 2443 (rs7273389), to 3338 (rs11634974), to 3342 (rs11636198 and rs12905212). For PheWASs in African ancestry individuals, all variants were tested for 1152 distinct phenotypes.

We found Bonferroni-corrected significant results for fifteen traits only in European individuals: rs11634974, rs11636198 and rs12905212 showed association with the same 6 traits, including ‘Current tobacco smoking’ which had the same direction of effect as in this study, and ‘Qualifications: College or University degree’ with opposite direction of effect (Supplementary Table 6). The fourth variant, rs7273389, showed nine significant associations, seven of which were with body fat measures (Supplementary Table 6).

## Discussion

Smoking is a preventable risk factor for several diseases worldwide^1^ with 80% of smokers living in low- and middle-income countries and a rising prevalence in Africa^3^. GWASs have shown that genetics plays a role in smoking behaviours^16,17^, but similarly to other traits, most studies have been performed in individuals of European ancestry, thus underestimating the role of genetic diversity for these traits globally^18,19^. Disentangling the genetics of smoking in sub-Saharan Africa is essential to shed light onto its biology in this region and globally, and to help elucidate its role as a risk factor for non-communicable diseases, either directly or through interaction^71^.

In this study we focussed on understanding the genetics of two smoking behaviour traits, smoking initiation and cessation, in 10,558 men of African ancestry living in five countries in the African continent and the UK, including participants from three cohorts: AWI-Gen^22^, divided into three geographical areas (East, South and West), UGR^23^, and UKB-AFR^24^. After a two-step meta-analysis, we identified 7 loci associated with SI and one with SC, all in tier2 (variants with a meta-analysis p-value < 5 × 10^−6^ and p-value < 0.05 in each cohort). We selected variants for *in silico* functional follow up analyses based on their posterior inclusion probability of being causal, obtaining 136 variants for SI and 36 for SC. We compared the allele frequencies at these variants between the African (AFR) and European (EUR) superpopulations from 1000GP^30^ obtained via VEP^51^. All but one variant of five associated loci with SI (on chr 1, 4, 7, 10 and 16) were monomorphic or had a MAF < 2% in EUR, while they were common (MAF ranging 2-25%) in AFR (Supplementary Table 7). Despite being common in both AFR and EUR, most variants on chr 15 showed allele frequencies 2-5 times higher in AFR, and the only variant for chr 19 was common for both groups (Supplementary Table 7). Variants in the chr 16 locus associated with SC showed a general higher frequency in EUR (Supplementary Table 7).

The variants identified by the few studies including African-American individuals described in the introduction^13, 14, 20^ did not replicate in our study, with the caveat that they focused on smoking phenotypes different from our traits.

Our gene prioritisation strategy highlighted *AC073133*.*1, MAN2B2*, and *SEMA6D* for SI and *CES1* and *LPCAT2* for SC, as genes supported by two out of the four analyses included (CADD score, eQTLs, pQTLs and chromatin interaction). A detailed description of their function, and of the genes highlighted by the pathway and protein-protein interaction analyses is included in Supplementary Note 3. *SEMA6D* on chr 15, however, will be described in detail here as this locus shows strong evidence of replication, is involved in one of the two pathways identified, and includes eQTLs for a gene involved in a protein-protein interaction (*FBN1*). This gene is a member of the semaphorin family that encodes both secreted and membrane proteins involved in axon guiding, that may have a role in maintaining and remodelling neuronal connections (see URLs). It was already identified as associated with smoking initiation, cessation and amount by five studies^17,21,44,45,46^ including the largest study to date by GSCAN^17^ and the recent trans-ethnic GWAS meta-analysis of smoking trajectories in the MVP cohort^21^. Querying GWAS Catalog for *SEMA6D* (as of July 2021), we found reported associations for 63 traits (see URLs), including smoking and drinking behaviour phenotypes, depression and cognitive ability (see URLs). In agreement with our PheWAS, educational attainment and body mass index phenotypes were among the top five trait classes associated with *SEMA6D* (Supplementary Table 6).

We are aware this study has its limitations. The underrepresentation of individuals of African ancestry in biobank-scale cohorts affects several aspects of this work: from the limited sample sizes to the availability of additional datasets for larger meta-analyses, replication and follow up analyses. Not only does this influence the number of datasets available for genomics studies, but also the number of variants that can be tested for association, as shown by half of the tier2 loci being monomorphic or rare in EUR from 1000GP^30^ (Supplementary Table 7). The sex bias observed in our datasets restricted the analyses to men only, impacting further on sample sizes and calling for specific attention to the cultural habits that may generate it. While it is true that smoking prevalence is very low among women in many African countries^10,11^, it is growing among girls^6^. The widespread use of chewing tobacco in some areas suggests that a new way of collecting data on tobacco use should be considered to aim to study nicotine dependence at a population level.

This study adds support to a previously identified locus from large European and trans-ethnic studies, but importantly it highlights the need for additional African cohorts to be developed and maintained. This is essential if we aim to overcome the limitations described above and be in a position to perform statistically powerful large-scale association studies across smoking behaviour phenotypes, as well as many other traits currently understudied in African populations.

## Data Availability

Complete summary statistics from the meta-analysis step2 for SI and SC are being deposited to NHGRI-EBI GWAS catalog [https://www.ebi.ac.uk/gwas/] (Study accession numbers: SI, GCST90091238; SC, GCST90091239; under embargo until publication is accepted).

## Data access statement

Individual-level genetic and phenotypic data from the AWI-Gen, Uganda Genome Resource and UK Biobank are available to approved researchers upon application or data access request.

File handling and individual analyses were performed using a combination of bash and R scripts, available upon request from the authors.

## Ethics approvals

The AWI-Gen study was approved by the Human Research Ethics Committee (Medical) of the University of the Witwatersrand (Wits) (protocol numbers M121029 and M170880). In addition, each research site obtained approval from their local ethics review board prior to commencing any participant-related activities. Uganda Genome Resource was approved by the Science and Ethics Committee of the UVRI, the Ugandan National Council for Science and Technology, and the East of England-Cambridge South NHS Research Ethics Committee United Kingdom. This research has been conducted using the UK Biobank Resource under approved Application 4892. Informed consent was obtained from all participants and all research was performed in accordance with relevant guidelines and regulations.

## Author contributions

Study design: MDT, MR, SF, AC, CB; Data collection: PRB, MR, SF, AC; Data preparation and initial processing: N-NP, CS, NS, J-TB, AC, CB; Data analyses: N-NP, PRB, CS, NS, KC, J-TB, CB; Manuscript writing: N-NP, CB with contributions from all authors. All authors critically evaluated and approved the manuscript.

MDT, MR, SF, AC and CB designed the study. PRB, MR, SF and AC collected the data. N-NP, CS, NS, J-TB, AC and CB prepared the data for analyses and performed initial quality controls. N-NP, PRB, CS, NS, KC, J-TB and CB performed data analyses. N-NP and CB wrote the manuscripts and prepared figures and tables, with contributions from all authors. All authors critically evaluated and approved the manuscript.

## Acknowledgements

We thank all volunteers from the AWI-Gen, UGR and UKB studies who have made this project possible. The AWI-Gen Collaborative Centre is funded by the National Human Genome Research Institute (NHGRI), Office of the Director (OD), Eunice Kennedy Shriver National Institute Of Child Health & Human Development (NICHD), the National Institute of Environmental Health Sciences (NIEHS), the Office of AIDS Research (OAR) and the National Institute of Diabetes and Digestive and Kidney Diseases (NIDDK), of the National Institutes of Health under award number U54HG006938 and its supplements, as part of the H3Africa Consortium. This research has been conducted using the UK Biobank Resource under Application 4892. This work was supported by the University of Leicester and Health Data Research UK, an initiative funded by UK Research and Innovation, Department of Health and Social Care (England) and the devolved administrations, and leading medical research charities. This study used the ALICE and SPECTRE High Performance Computing Facilities at the University of Leicester and the ZA-Wits-Core Cluster at University of the Witwatersrand. As this research was funded in whole, or in part, by the Wellcome Trust, for the purpose of open access, the author will apply a CC BY public copyright licence to any Author Accepted Manuscript version arising from this submission.

C.B. was supported by an internal fellowship at the University of Leicester from the Wellcome Trust Institutional Strategic Support Fund (204801/Z/16/Z) and a UKRI Innovation Fellowship at Health Data Research UK (MR/S003762/1). M.D.T. is supported by a Wellcome Trust Investigator Award (WT202849/Z/16/Z). M.R. is a South African Research Chair in Genomics and Bioinformatics of African populations hosted by the University of the Witwatersrand, funded by the Department of Science and Technology, and administered by the National Research Foundation. C.S. acknowledges H3Africa Bioinformatics Network (H3ABioNet) Node, National Biotechnology Development Agency (NABDA), and the Center for Genomics Research and Innovation (CGRI) Abuja, Nigeria. S.F. is funded by the Wellcome International Intermediate fellowship (220740/Z/20/Z) at the MRC/UVRI and LSHTM.

The views expressed here do not necessarily reflect the views of the funders.

## URLs

GWASCatalog: https://www.ebi.ac.uk/gwas/api/search/downloads/alternative;

eQTLs data sources in FUMA: https://fuma.ctglab.nl/tutorial#eQTLs;

Chromatin Interaction datasets in FUMA: https://fuma.ctglab.nl/tutorial#chromatin-interactions;

BRAINEAC: http://www.braineac.org/;

IEU OpenGWAS project: https://gwas.mrcieu.ac.uk/datasets/;

ukb-b: http://www.nealelab.is/blog/2017/7/19/rapid-gwas-of-thousands-of-phenotypes-for-337000-samples-in-the-uk-biobank;

ukb-d: http://www.nealelab.is/uk-biobank;

ukb-e: https://pan.ukbb.broadinstitute.org/;

GeneCards - *SEMA6D*: https://www.genecards.org/cgi-bin/carddisp.pl?gene=SEMA6D;

GWASCatalog-*SEMA6D*: https://www.ebi.ac.uk/gwas/genes/SEMA6D

## Notes

### Competing Interest Statement

The authors have declared no competing interest.

### Funding Statement

Please refer to the acknowledgements section for details about funding support.

### Author Declarations

The AWI-Gen study was approved by the Human Research Ethics Committee (Medical) of the University of the Witwatersrand (Wits) (protocol numbers M121029 and M170880). In addition, each research site obtained approval from their local ethics review board prior to commencing any participant-related activities. Uganda Genome Resource was approved by the Science and Ethics Committee of the UVRI, the Ugandan National Council for Science and Technology, and the East of England-Cambridge South NHS Research Ethics Committee United Kingdom.

### Summary of Updates

Distribution and reuse options updated.

