## Supplementary Materials for "Genetic insights into smoking behaviours in 10,558 men of African ancestry from continental Africa and the UK"

### Supplementary Note 1

#### Definition of smoking behaviour traits from questionnaire data

##### AWI-Gen

The phenotype definitions combine two questions:

- 9.1.1: “Have you ever smoked any tobacco products such as cigarettes, cigars or pipes?”
- 9.1.2: “Do you currently smoke any tobacco products, such as cigarettes, cigars or pipes?”

Never smokers have never smoked any tobacco products [Q9.1.1 = no]. Current smokers smoke at present [Q9.1.2 = yes]. Previous smokers have smoked at some point of their life but do not smoke at present [Q9.1.1 = yes & Q9.1.2 = no]. Current and Previous smokers were combined into Ever smokers.

##### UGR

The phenotype definitions combine three questions:

- Tobac1: “Do you currently smoke any tobacco products, such as cigarettes, cigars or pipes?”
- Tobac2: “If you do currently smoke tobacco products (tobac1), do you currently smoke tobacco products daily?”
- Tobac3:”In the past, did you ever smoke daily?”

Never smokers do not smoke at present [Tobac1 = no] and have never smoked daily [Tobac3 = no]. Current smokers smoke at present [Tobac1 = yes]; this includes participants that smoke daily at present [Tobac2 = yes], smoked daily in the past [Tobac3 = yes] or never smoked daily [Tobac2 = no & Tobac3 = no]. Previous smokers do not smoke at present [Tobac1 = no] but smoked daily in the past [Tobac3 = yes]. Current and Previous smokers were combined into Ever smokers.

##### UKB-AFR

The phenotype definitions combine three questions:

- 1239: “Do you smoke tobacco now?”
- 1249: “In the past, how often have you smoked tobacco?”
- 2644: “In your lifetime, have you smoked a total of at least 100 times?”

Never smokers are those individuals who do not smoke at present and never smoked in the past [1239=0 & 1249=4] or do not smoke at present, smoked occasionally or just tried once or twice in the past, but had less than 100 smokes in their lifetime [1239=0 &1249=2/3 & 2644=0].
Previous smokers do not smoke at present and smoked on most or all days in the past [1239=0 &1249=1] or do not smoke at present, smoked occasionally or just tried once or twice in the past, and had more than 100 smokes in their lifetime [1239=0 &1249=2/3 & 2644=1]. Current smokers smoke at present, on most or all days or occasionally [1239=1/2]. When combining Previous and Current into Ever smokers, we include also individuals who smoked on most/all days or occasionally, had more than 100 smokes, in the past but prefer not to answer about current smoking [1239=-3 & 1249=1 or 1239=-3 & 1249=2 & 2644=1].

### Supplementary Note 2

#### Discovery analyses: genome-wide association testing in each individual cohort and meta-analysis step1

We performed a genome-wide association analysis for each dataset and phenotype combination in a total of 10,558 men for SI and 4,257 for SC (Supplementary Figure 1). For the SI trait, we observed variants with p-value below the suggestive threshold at $5 \times 10^{-6}$ for all datasets, with one variant reaching genome-wide significance at $5 \times 10^{-8}$ in either AWI-west (rs74326809) or UGR (rs114033989) (Supplementary Figure 5). The qq-plots suggested a good correction for population stratification and relatedness (Supplementary Figure 5), and in agreement with this the LD score intercept values were all below 1.05 (Supplementary Table 8). For SC, we observed variants with p-value below the suggestive threshold at $5 \times 10^{-6}$ for all datasets, with at least one variant reaching genome-wide significance in all three AWI-Gen datasets (AWI-East: one variant [rs559053]; AWI-South: 11 variants on two loci [rs10756181, rs7858303, rs10756185, rs2382262, rs2382263, rs2382264, rs2382265, rs531269074, rs189304958, rs10420605 and rs13343613]; AWI-West: one variant [rs111929469]; Supplementary Figure 6). The qq-plots showed good correction for population structure (Supplementary Figure 6). In this case, the LD score intercept for all datasets was below 1.05 with the exception of AWI-south (1.08) which required genomic correction on the standard errors and recalculation of the p-values (Supplementary Table 8).

A modified random effect model was implemented for both steps of the meta-analysis on variants present in at least two of the individual datasets. Step1 included the three AWI-Gen datasets (Supplementary Figure 1). For SI, step1 included 12,960,953 variants: no SNPs passed the genome-wide significant threshold while 33 variants were discovered at the suggestive threshold (Supplementary Figure 7a). The qq-plot showed a good correction for population structure (Supplementary Figure 7a) and the LD-score regression intercept was 0.98. For SC, step1 analysed a total of 12,034,951 SNPs: no genome-wide significant variants were observed and 48 variants had p-values below $5 \times 10^{-6}$ (Supplementary Figure 7b). The qq-plot confirmed that we applied a good correction for population structure (Supplementary Figure 7b) and the LD-score regression intercept was 0.95.

### Supplementary Note 3

#### Description of the genes identified in the study

Our gene prioritisation strategy highlights *AC073133.1*, *MAN2B2*, and *SEMA6D* for SI and *CES1* and *LPCAT2* for SC, as genes supported by two out of the four analyses included (CADD score, eQTLs, pQTLs and chromatin interaction). *SEMA6D* is described in detail in the main text.

*AC073133.1* codifies for a long intergenic non-coding RNA, which is a biotype involved in different activities such as chromatin remodelling or transcription regulation, probably influencing the expression of other genes and having tissue specificity[1]. *MAN2B2* is a protein coding gene on chr4 for a mannosidase (see URLs). *CES1* and *LPCAT2* are both protein coding genes and they are associated with different variants for both eQTLs and chromatin interaction. *CES1* encodes for a carboxylesterase involved in xenobiotics metabolism, playing a major role in drug clearance in the liver (see URLs); *LPCAT2* is a lysophospholipid acyltransferase (see URLs).

Gene pathway analysis was performed only on the 55 genes we could query through an entrez ID and no information was retrieved for the remaining identified genes for SI; no pathway analysis was performed for SC as there was only one locus associated for this phenotype. The two pathways identified by this analysis highlighted two groups of genes: (i) *FBN1*, *WFS1* and *SHH* and (ii) *SHH*, *PDPN*, *SALL1* and *SEMA6D* (Supplementary Table 5).
*FBN1* encodes for a protein in the fibrillin family involved in the force-bearing activity of connective tissues (see URLs). Mutations in this gene are associated with several chronic diseases (see URLs). In our analysis, *FBN1* shows eQTL results in the putamen region of the brain for three variants on chr 15, among which the sentinel variant for this associated locus (Supplementary Table 4a). *WFS1* regulates beta-cell activity and mutations in this gene are associated with the Wolfram syndrome (see URLs). Sandhu et al. have found that common variations in *WFS1* confers risk for type 2 diabetes in UK and Ashkenaz populations[2]. In our study, *WFS1* shows chromatin interaction results in both fetal and adult cortex tissues with four variants in chr 4 including the sentinel variant for this associated locus (Supplementary Table 4a). *SHH* encodes for a protein that has a key role in early embryonic development and mutations in this gene can cause deformities in several parts of the body, in particular the forebrain (see URLs). Ten variants on chr 7 show evidence of chromatin interaction with *SHH* found in a cell line of lung fibroblasts and neural progenitor cells (Supplementary Table 4a).
*PDPN* produces a glycoprotein expressed in several human tissues whose function is yet to be determined and it has been suggested as a marker of lung injury (see URLs). Five variants on chr 1 show chromatin interaction results with *PDPN* in lung fibroblast cells (Supplementary Table 4a). *SALL1* encodes for a transcriptional repressor for organogenesis with mutations in this gene causing the Townes-Brocks syndrome and the bronchio-oto-renal syndrome (see URLs). In our analysis, *SALL1* shows evidence of chromatin interaction with four variants on chr 16 found in lung fibroblast cells (Supplementary Table 4a).

Regarding the protein-protein interactions, *CYP4F3* and *CYP4F8* encode two monooxygenases belonging to the cytochrome P450 family involved in drug metabolism and lipids synthesis (see URLs). No association with smoking behaviour has been found previously with either of these genes, but the activity of enzymes of the same family has been shown to be induced by tobacco smoke, specifically *CYP1A2*, *CYP2A6* and *CYP2B6*[3,4,5,6,7]. The protein-protein interaction between *FBN1* and *WFS1* is supported by two levels of evidence: they are both enriched for the same pathway ('metanephros development') and they show chromatin interaction for brain tissue. In addition, Wu et al. performed a gene-environment-wide interaction study (GEWIS) in order to discover any gene-by-smoking interaction for type 2 diabetes or fasting glucose in Europeans and African Americans[8]. Interestingly, *FBN1* was discovered as associated with lower risk of type 2 diabetes in African American smokers only, through the variant rs140637[8] which shows the same direction of effect of 2 out of three variants associated with SI in our study and mapped to *FBN1*. Opposite to *FBN1*, mutations in *WFS1* are known to increase the risk of type 2 diabetes[2] showing the need to improve our knowledge about the implication of these two genes in the context of type 2 diabetes.

**URLs**

GeneCards - *MAN2B2*: https://www.genecards.org/cgi-bin/carddisp.pl?gene=MAN2B2;
GWASCatalog - *MAN2B2*: https://www.ebi.ac.uk/gwas/genes/MAN2B2;
GeneCards - *CES1*: https://www.genecards.org/cgi-bin/carddisp.pl?gene=CES1;
GeneCards - *LPCAT2*: https://www.genecards.org/cgi-bin/carddisp.pl?gene=LPCAT2;
GeneCards - *FBN1*: https://www.genecards.org/cgi-bin/carddisp.pl?gene=FBN1;
GeneCards - *SHH*: https://www.genecards.org/cgi-bin/carddisp.pl?gene=SHH;
GeneCards - *WFS1*: https://www.genecards.org/cgi-bin/carddisp.pl?gene=WFS1;
GeneCards - *PDPN*: https://www.genecards.org/cgi-bin/carddisp.pl?gene=PDPN;
GeneCards - *SALL1*: https://www.genecards.org/cgi-bin/carddisp.pl?gene=SALL1;
GeneCards - *CYP4F3*: https://www.genecards.org/cgi-bin/carddisp.pl?gene=CYP4F3;
GeneCards - *CYP4F8*: https://www.genecards.org/cgi-bin/carddisp.pl?gene=CYP4F8;

### Supplementary Figures

#### Supplementary Figure 1:

Diagram of overall study design. SI, smoking initiation; SC, smoking cessation; AWI, Africa Wits-INDEPTH Partnership for Genomic Studies; UGR, Uganda Genome Resource; UKB-AFR, individuals of African ancestry in UK Biobank; MAC, minor allele count; MAF, minor allele frequency; PIP, posterior inclusion probability; D' and $r^2$, measures of linkage disequilibrium.

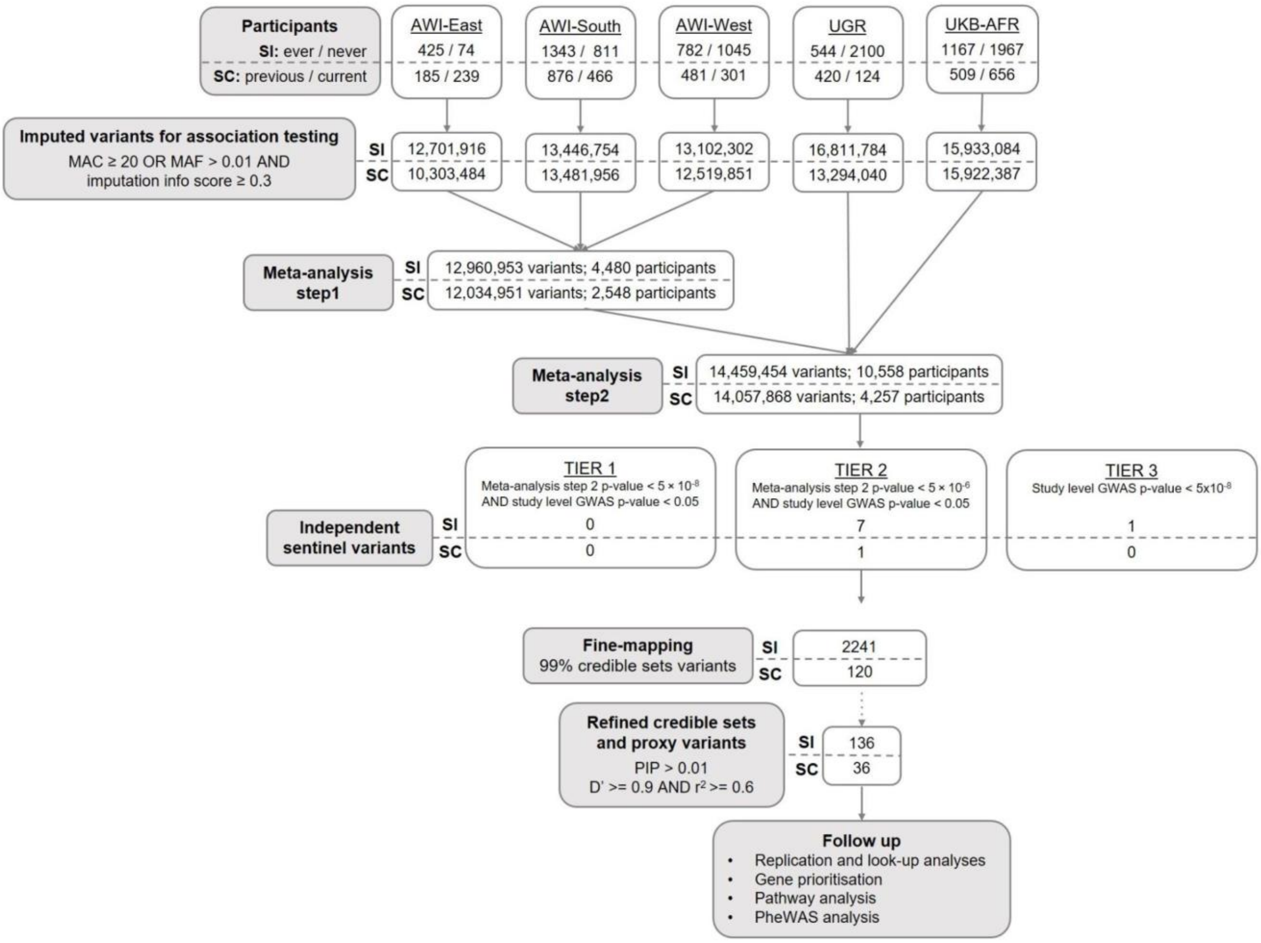

**Supplementary Figure 2:**

Q-Q plots of meta-analysis step 2. a) smoking initiation; b) smoking cessation. Number of participants and variants analysed is reported in Supplementary Figure 1.

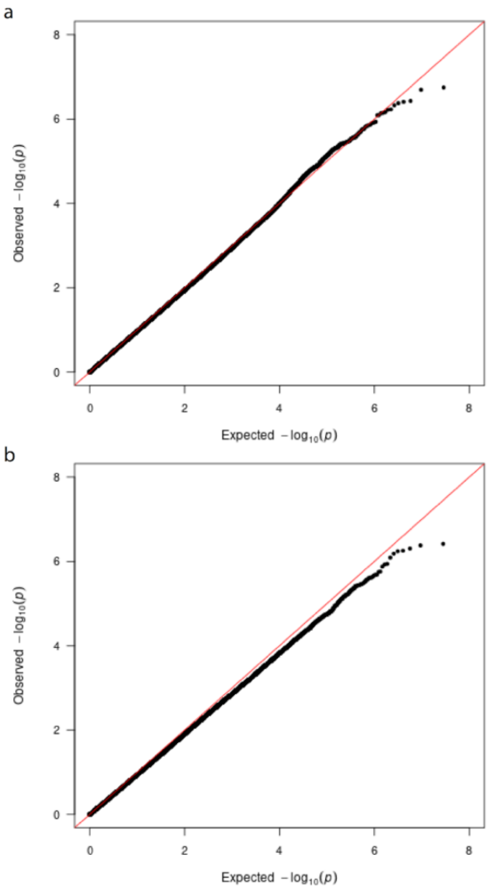

#### **Supplementary Figure 3:**

Forest plot of sentinel variants for SI and SC. The variants are reported in the following format: Chromosome:Position(b37)_non effect allele_effect allele; OR, odds ratio; 95% CI, 95% confidence interval; AWI-Gen, Meta-analysis step1; UGR, Uganda Genome Resource; UKB-AFR, individuals of African ancestry in UK Biobank. The dotted red line represents OR = 1.

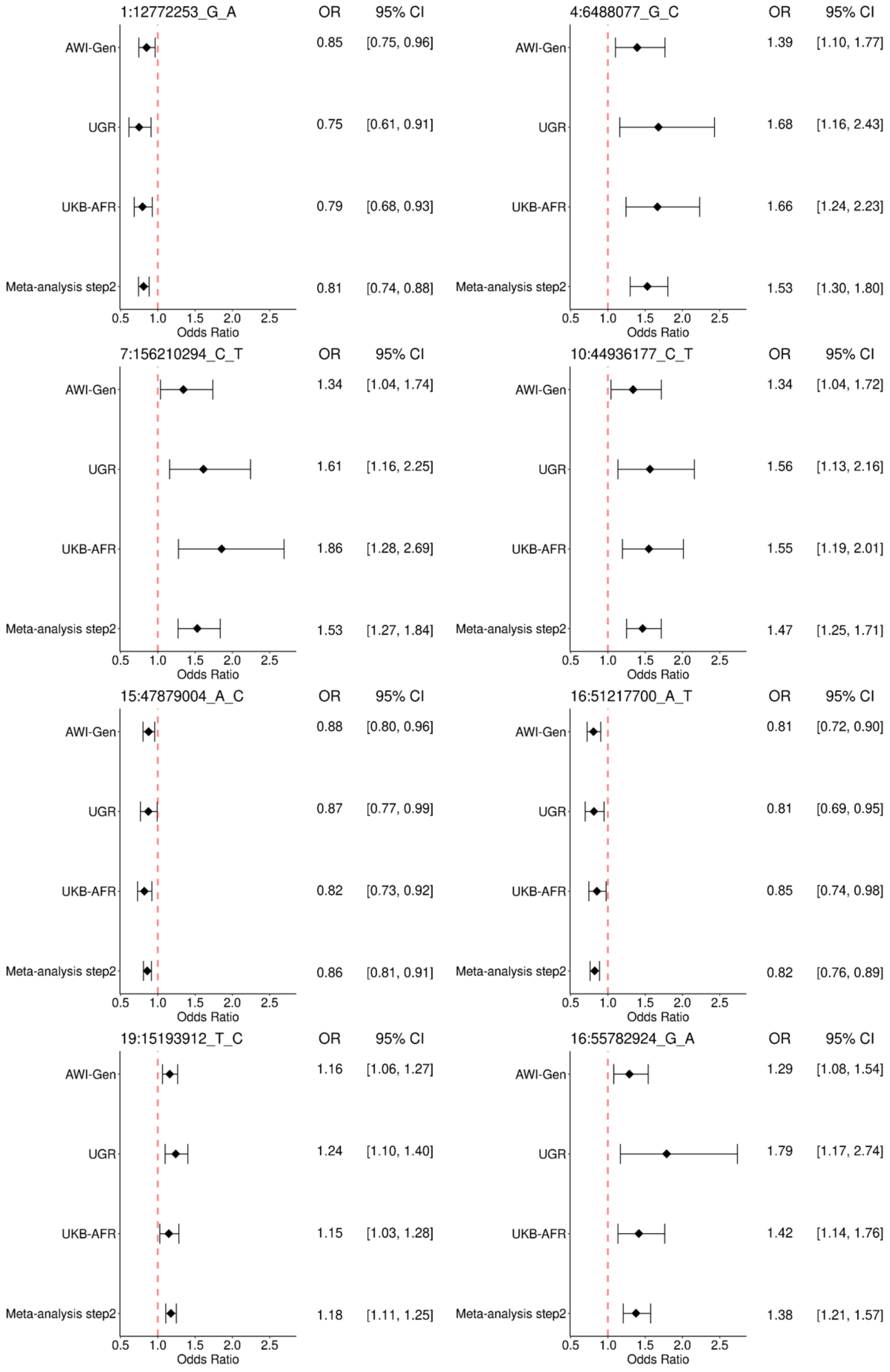

#### Supplementary Figure 4:

Locus zoom plot of sentinel variants. For smoking initiation: a) chr1: rs116934871; b) chr4: rs114828540; c) chr7: rs75104774; d) chr10: rs10160111; e) chr15: rs11636198; f) chr16: rs74019935; g) chr19: rs4808946. For smoking cessation: h) chr16: rs12443715. Sentinel variants are represented by a diamond shape; $r^2$, measure of linkage disequilibrium. Genes located in a region of +/- 100 kb from the sentinel variant (b37 position) are reported at the bottom of each plot.

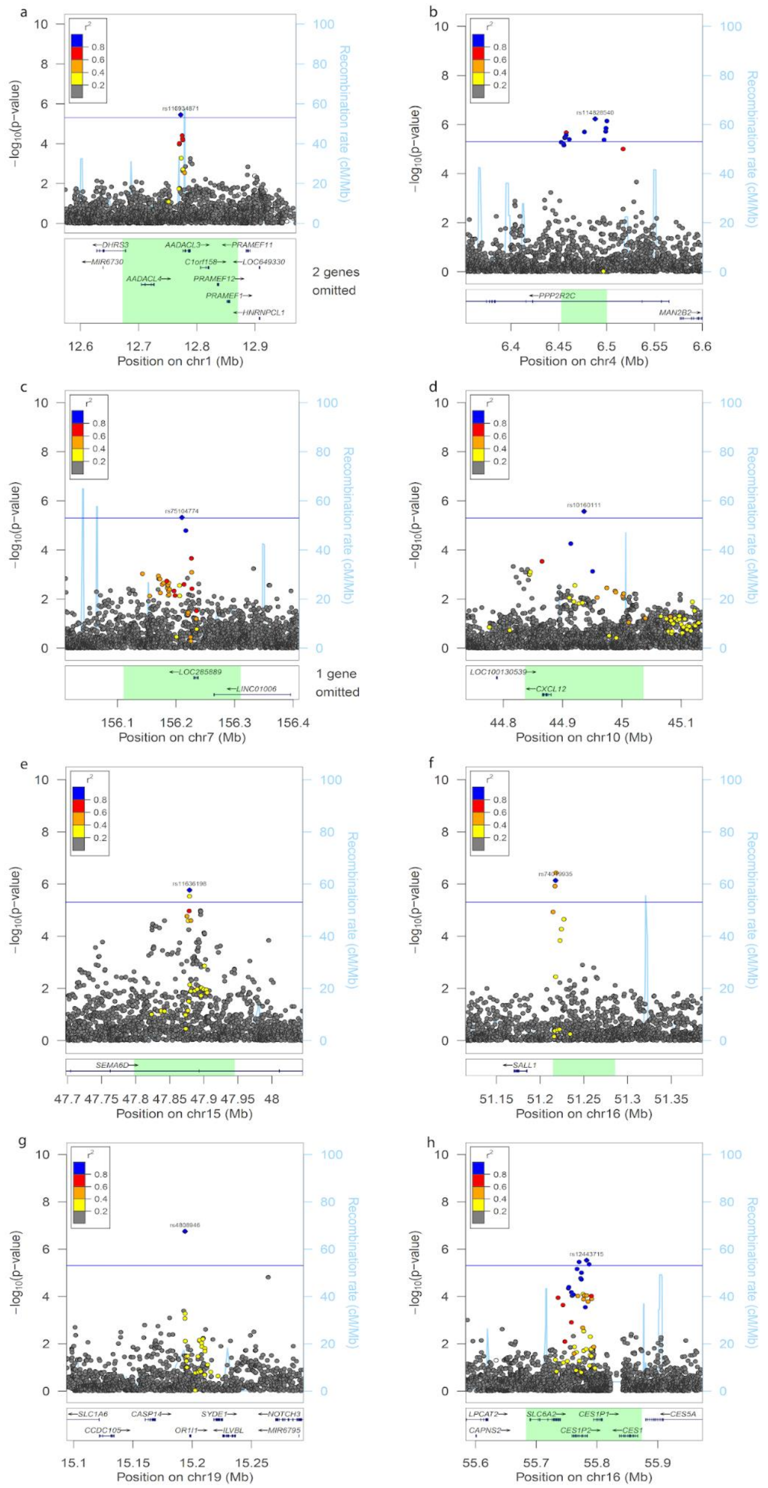

#### Supplementary Figure 5:

Manhattan and qq- plots of SI study level GWAS. a) AWI-East; b) AWI-South; c) AWI-West; d) UGR; e) UKB-AFR. Manhattan plot: continuous line is the suggestive p-value significance threshold (5 x $10^{-6}$) and the dashed line is the genome-wide significance threshold (5 x $10^{-8}$). Number of participants and variants analysed is reported in Supplementary Figure 1. AWI, Africa Wits-INDEPTH Partnership for Genomic Studies; UGR, Uganda Genome Resource; UKB-AFR, individuals of African ancestry in UK Biobank.

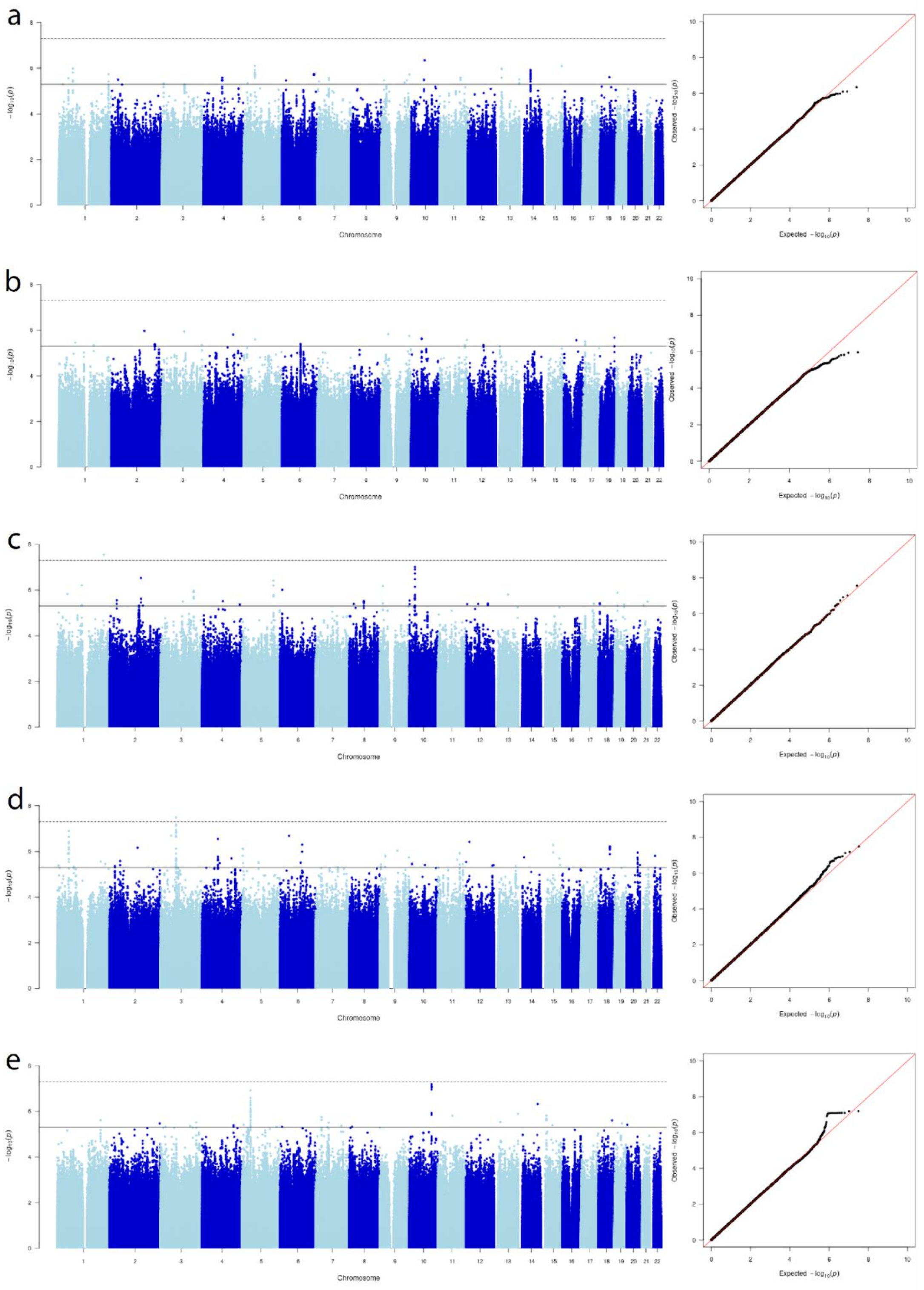

#### Supplementary Figure 6:

Manhattan and qq- plots of SC study level GWAS. a) AWI-East; b) AWI-South; c) AWI-West; d) UGR; e) UKB-AFR. Manhattan plot: continuous line is the suggestive p-value significance threshold (5 x $10^{-6}$) and the dashed line is the genome-wide significance threshold (5 x $10^{-8}$). Number of participants and variants analysed is reported in Supplementary Figure 1. AWI, Africa Wits-INDEPTH Partnership for Genomic Studies; UGR, Uganda Genome Resource; UKB-AFR, individuals of African ancestry in UK Biobank.

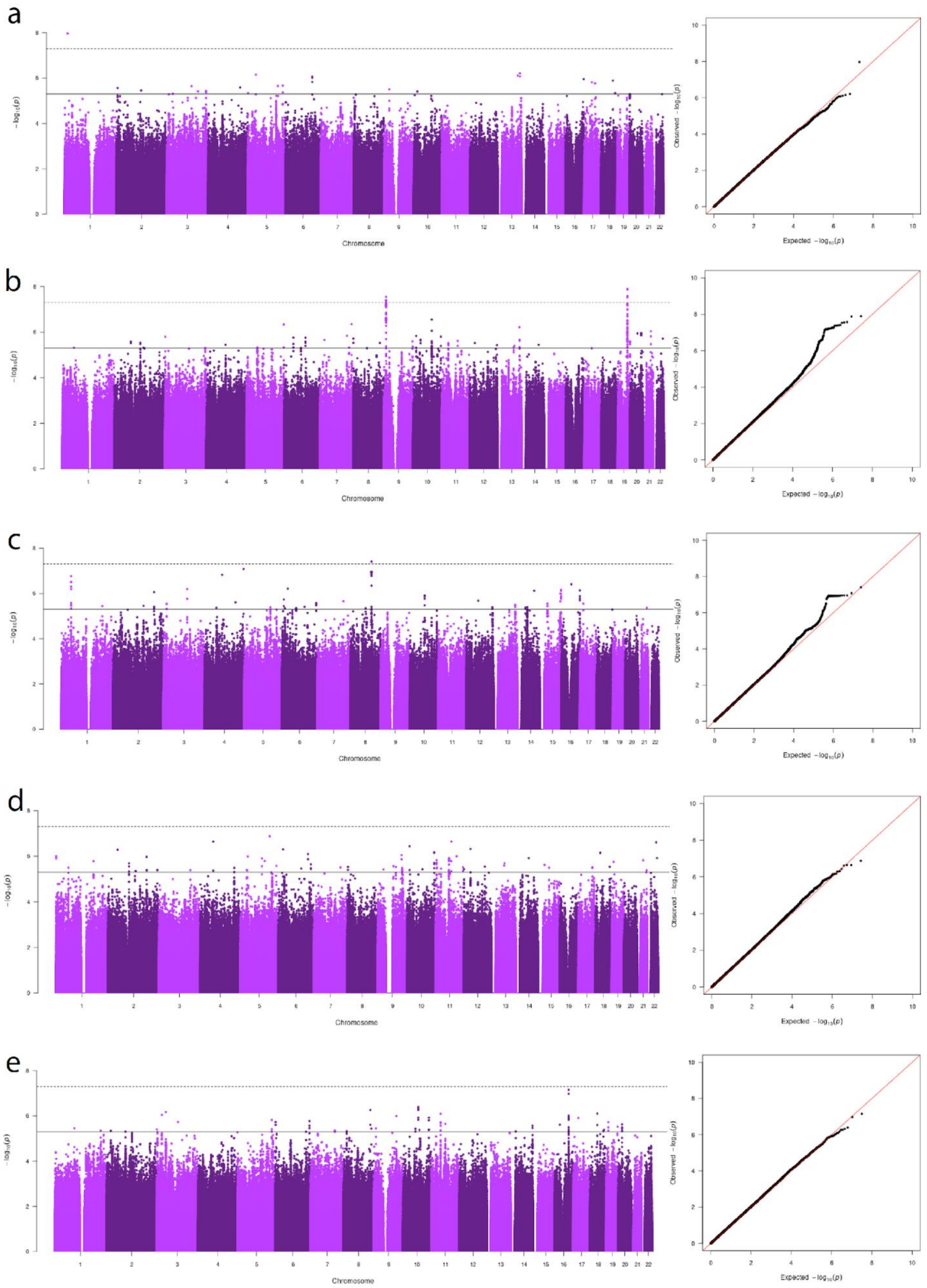

#### Supplementary Figure 7:

Manhattan and qq- plots of meta-analysis step 1. a) smoking initiation; b) smoking cessation. Number of participants and variants analysed is reported in Supplementary Figure 1.

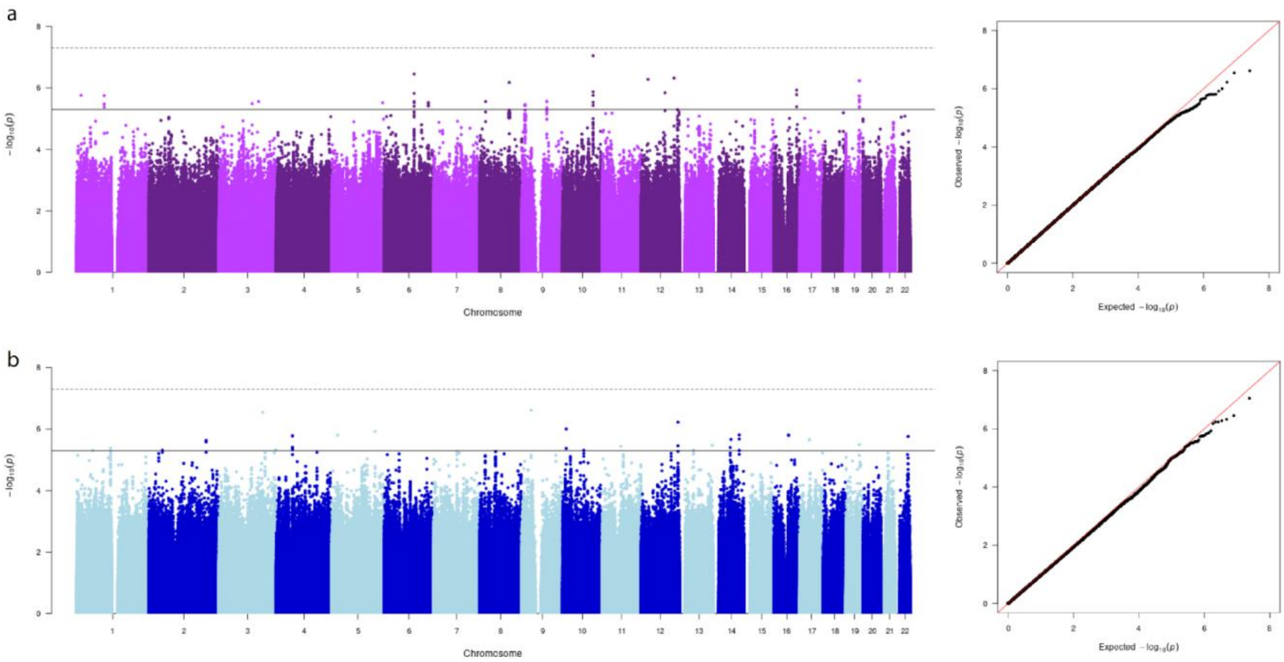
